# Understanding preferences for self-sampling in a national cervical screening programme: a protocol for a discrete choice experiment

**DOI:** 10.1101/2025.02.12.25322143

**Authors:** Shabnam Thapa, Jennifer C. Davies, Emma J. Crosbie, Katherine Payne, Stuart J. Wright

**Affiliations:** National Institute for Health and Care Excellence, Manchester, UK; Manchester Centre for Health Economics, The University of Manchester, Manchester, UK; Gynaecological Oncology Research Group, Division of Cancer Sciences, Faculty of Biology, Medicine and Health, University of Manchester, Manchester, UK; Department of Obstetrics and Gynaecology, Manchester Academic Health Science Centre, St Mary’s Hospital, Manchester University NHS Foundation Trust, Manchester, UK

## Abstract

**Introduction:** The NHS cervical screening programme (NHSCSP) currently involves a healthcare professional collecting a cervical sample in a healthcare setting. This method of screening has barriers associated with access to screening appointments, and the poor acceptability of the speculum examination. Primary screening through HPV testing has led to the development of self-sampling screening methods including vaginal and urine self-sampling, with many UK studies comparing these screening methods with the current NHSCSP. It is not known what features of self-sampling influence individual’s preferences and cervical screening uptake. To understand these preferences, we plan to undertake a discrete choice experiment (DCE). This protocol aims to describe the steps taken to design the DCE and the proposed approach to fielding the DCE to identify preferences for different sampling approaches in cervical screening.

**Methods and analysis:** An online survey comprising a DCE was designed to understand preferences of individuals for self-sampling methods within the NHSCSP. Attributes and levels for the DCE were generated through an iterative process including a literature review of qualitative studies about self-sampling cervical screening methods, input from cervical screening clinical experts and a patient and public involvement group (n=6). A D-efficient design was used to create choice sets for the DCE survey. Regression based analysis will be used to estimate the impact of each attribute and level on individual choices.

**Ethics and dissemination:** This study has been approved by The University of Manchester Proportionate Research Ethics Committee (2024-20767-37669). The results of the DCE will be submitted for publication in a relevant peer review journal and the results will be presented at national and international conferences.

**STRENGTHS AND LIMITATIONS OF THIS STUDY:**

- To our knowledge, this study will be the first to quantify stated preferences for self-sampling methods within a CSP focusing on all individuals who are eligible for screening in the UK.
- The attributes and levels will be selected based on previous qualitative work as well as from multiple discussions with relevant stakeholders, including clinicians and public contributors.
- The results will contribute towards understanding what aspects of self-sampling methods within a CSP are important for the target population.
- Since the survey will only be available online, there is a potential to miss individuals without technological access and literacy.

## INTRODUCTION

Cervical cancer is one of the most common cancers affecting women and people with a cervix worldwide, with approximately 660,000 new cases annually (1).⍰ In the UK, around 3,200 women are diagnosed with cervical cancer each year, with the highest incidence observed amongst individuals aged 30 to 34 years (2). Cervical cancer is caused by persistent infection with high-risk human papillomavirus (HPV) (3,4). HPV is a common virus which is responsible for around 99% of cervical cancer cases (5).

The current national cervical screening programme (CSP) in the UK targets people with a cervix aged between 25 to 64 years, with a total of 3.43 million attending in 2022-23 (6). Those registered with a GP are invited for screening every 3 or 5 years, depending on age and location within the nations of the UK. At present, screening involves a healthcare professional performing a speculum examination to obtain a cervical sample in a healthcare setting. Since 2019, cervical screening utilises primary HPV testing, where the cervical sample is first examined for the presence of the HPV virus. If the sample if HPV negative, no further tests are required, and the individual returns to routine recall and will be offered screening in 3 or 5 years. If the result is HPV positive, cytology is performed to triage the individual for further investigations. Depending on the cytology results, the individual may be invited for another screen in a year or referred for a colposcopy if abnormal cells are detected (7).

Since the introduction of an organised cervical screening programme in England in 1988, cervical cancer deaths have reduced by approximately 70% (8). Despite this the uptake of cervical screening is declining, with only 68.7% of eligible individuals screened in 2022/2023, a decrease of 1.2% from the previous year (5).

Several barriers to attendance for cervical screening have been identified, including poor access to screening appointments, lack of time to attend screening, poor acceptability of the speculum examination, fear of cancer, low perceived risk, and discomfort with the gender of the healthcare professional performing the sampling (9,10). Uptake is particularly low among individuals from ethnically diverse backgrounds, those younger than 30 years old, people from socio-economically deprived communities and those who identify as LGBTQ+ (11,12).

To overcome these barriers and increase uptake, there has been growing interest in alternative cervical screening sampling methods such as a self-collected vaginal swab and urine samples. Several countries, including the Netherlands, Malaysia, and Albania, offer home based self-sampling as a primary screening option (13–15). Other countries such as Australia, Denmark, Finland, and France, offer self-sampling to under screened populations (16). UK based studies are currently being conducted to compare the test performance of self-sampling with healthcare practitioner taken cervical sampling (17–19). In clinical trials, such as the UK based HPValidate trial, vaginal self-sampling has been shown to have similar diagnostic accuracy to cervical sampling (20,21). However recent real-world prospective data from the Netherlands, where vaginal self-sampling has been a whole population opt-in option since 2017, shows a decrease in HPV positivity and CIN3+ rates in self-sampling compared to healthcare practitioner-taken cervical sampling which has raised some concern around its comparative test accuracy (22,23). Urine self-sampling also shows promising diagnostic test accuracy compared to healthcare practitioner taken cervical sampling if collected with a specialised first void urine collection device (24,25). Its high acceptability rate (26) may potentially improve cervical screening uptake further.

The full potential of cervical screening will only be achieved if there is sufficient uptake. While self-sampling is likely to increase uptake by overcoming some of the barriers of access to cervical screening, the improved uptake would need to be sufficient to offset any decrease in test sensitivity (27). Uptake of any service depends significantly on individual preference. It is yet to be understood what characteristics of sampling drive individual choice to attend screening or not. To explore individual preference for self-sampling methods for cervical screening, a discrete choice experiment (DCE) will be conducted.

A Discrete Choice Experiment (DCE) is a preference elicitation method used to generate preferences for characteristics (attributes) of an intervention by observing choices made to structured questions (28,29). In a DCE, individuals are presented with a series of hypothetical scenarios and asked to choose between different options. These hypothetical scenarios are described by key features of the intervention, known as attributes, and each attribute can have a range of values called levels. The responses to the DCE questions are then analysed to determine preferences, how attributes trade-off against each other, and the relative importance of each attribute (30). The results from the DCE can be used to predict the potential uptake for different sampling approaches for cervical screening. Heterogeneity in preferences can also be explored, informing the design of a future CSP delivery model that meets the need of the whole population and maximise its uptake and overall impact.

This paper aims to describe the steps taken to design a DCE to understand the preferences of people with a cervix for different attributes of a self-sampling approach within a CSP.

## METHODS AND ANALYSIS

### Overview of approach and methods

An online survey containing a DCE will be designed to elicit preferences from individuals eligible for the CSP. The survey will be developed using guidance for designing preference studies and subsequently reported in-line with recently published reporting guidelines (31,32).

In each DCE task the participant will be shown two unlabelled alternatives describing potential ways in which a sample could be taken for cervical screening. They will be asked to choose which of the alternatives they would prefer if they had to choose or whether they instead would choose to receive no screening. Participants will be asked to complete a series of these questions and the resulting preference data for all participants will be analysed using econometric models using logistic regression. The results will show which attributes have the biggest impact on driving the decision about whether to have screening and will allow the uptake for specific HPV sampling methods to be predicted.

### Ethical Approval

Proportionate Ethical approval for this study has been granted by The University of Manchester Research Ethics Committee. A participant information sheet will be shown on the first accessed digital page of the study platform and participants will be asked for consent to take part in the study. The participant information sheet will inform participants that the data from this study will be made publicly available upon submission of the work to a scientific journal. The survey will not explicitly record any personal information about participants and any free text comments will be removed before the data is stored in an open access repository.

### Defining the list of attributes and levels

Discrete choice experiments are underpinned by the theory that when an individual chooses between the available alternatives, they do this based on the attributes of the good or service on offer (33). The alternatives represent different hypothetical services that can be chosen by the participant. In this study, the alternatives will be different potential ways of taking the sample for cervical screening. These will be unlabelled which means they will be described to participants as sampling method 1, sampling method 2 and so on. The alternative would be to use a labelled approach where the alternatives would be labelled as specific sampling approaches, for example healthcare-practitioner obtained cervical sampling, vaginal self-sampling or urine self-sampling. Our approach was chosen because using a labelled approach would make it impossible to determine the value of some attributes that are directly related to the sampling approach. For example, healthcare-practitioner obtained cervical sampling can never be done in a home setting so it would not be possible to separate the value of self-sampling as an intervention from the value of being able to collect the sample at home.

Identifying the key attributes and levels of a service is critical to understanding how individuals choose between alternatives. The final list of attributes included in this DCE survey was developed through an iterative process, incorporating information from both published literature and relevant stakeholder input, which follows recommended practice when designing a DCE (34).

Initially, a list of potential attributes was identified by reviewing qualitative studies on the facilitators, barriers, acceptability, attitudes, and perceptions related to various methods of self-sampling methods within a CSP. A systematic approach to searching the literature was conducted in Medline and Embase (up to January 2024) to identify relevant studies. The full search strategy is available in the supplementary material. Google Scholar was used to identify any other relevant studies not found in primary databases. This review focused on studies conducted in the UK due to their direct relevance to the decision problem. Four UK based studies on self-sampling were identified (9,35–37). Given the limited number of qualitative studies specific to self-sampling methods in the UK, we expanded our search to include studies from other countries with similar socio-economic backgrounds (high income countries as defined by the World Bank) with universal health coverage, ensuring that screening methods are covered by national health systems without additional cost to the users. Exclusion criteria included studies that were not qualitative, did not involve individuals eligible for cervical screening, focused on health provider supply of self-sampling, were unpublished, or not available in English. A total of 17 relevant studies were identified, generating a list of 39 potential attributes (see supplementary material, Table 1).

The first step in refining these attributes was to screen them to ensure only those relevant to the decision problem were included. For this initial screening, we used a behavioural screening model, the Integrated Screening Action Model (I-SAM model), as a framework to categorise the attributes according to the seven stages of the screening process: unaware, unengaged, undecided, decided to act, acting, repeat and decided not to act (38). The I-SAM model was used as a framework for attribute selection as the decision to undergo screening involves a complex series of stages with an extensive range of influencing factors. Using the I-SAM model helped to distinguish attributes into those that may be used to make decisions at different stages and factors that might reflect participants’ attitudes, which influence their screening decisions. For example, perception of self-sampling methods might be shaped by education or cultural norms, while motivational factors, such as an understanding of HPV and cervical cancer, can encourage screening. Conversely, physical disabilities might discourage the use of self-sampling methods. While these factors are important, they do not necessarily function as characteristics of the cervical screening methods themselves. As a result of screening using the I-SAM model, a list of 14 potential attributes were kept. These included: sensitivity of the test; chance of recall; location of the test; method of the test; feeling of discomfort or pain; time taken to receive the results; how to organise an appointment; packaging of the kit; size of the brush; instructions provided with the kit; how the sample is returned; extended time frame of receiving final results for self-sampling; location of self-sampling.

This list of attributes would be too cognitively burdensome to include in a single DCE choice task (39,40), with the ideal number of attributes being around six (41). However, having too few attributes might also result in excluding a potentially important aspect of the alternative screening methods. The list of attributes was further refined only to include the most important attributes in the DCE survey. To facilitate this refinement, we sought input from relevant stakeholders to reduce the list of attributes and to identify any additional attributes that may have been overlooked. Our stakeholders included: i) clinical experts with expertise in cervical screening including self-sampling methods, and ii) a patient and public involvement (PPI) group comprising six individuals aged 25 to 64 years who had previously been invited for cervical screening, representing a diverse range of ethnic backgrounds and included individuals from other groups who face additional barriers in accessing screening, including those with physical disabilities and neurodivergence.

When the initial list of attributes was presented to clinical experts, they didn’t deem ‘chance of recall’ and ‘time to result’, to be factors that would be used to make a decision about screening method. Hence, these attributes were removed. They also provided feedback on the wording used to describe the attributes. In the PPI group meeting, members were asked their opinion on self-sampling methods and what factors might be important while making choices for cervical screening sampling type. From the PPI group meeting, a new attribute, ‘frequency of screening’ was suggested and subsequently added. This was because one member suggested they may be willing to have more frequent self-sampling if it could offset the reduced test sensitivity. The levels of the attributes were set with input from the clinical experts. The likelihood of detecting cervical abnormalities was set to cover the range of potential sensitivities for different types of self-sampling (22). The potential intervals for sampling were aligned to different intervals used in the NHSCSP for HPV positive individuals (1 year) or HPV negative individuals (3 or 5 years). 2-year and one-year intervals were added to represent the potential to shorten intervals to offset the reduced sensitivity of self-sampling. The final list of levels and attributes is presented in Table 1.

**Table 1.** Final list of attributes and levels

| Attributes | Description | Levels | Attribute Coding |
| --- | --- | --- | --- |
| Likelihood the sampling method detects cervical abnormalities if present | How accurate is the test to detect abnormalities in cervical cells? | 1. 80%<br>2. 85%<br>3. 90%<br>4. 95% | Continuous |
| How is the sample taken? | Process of taking the sample | 1. Healthcare practitioner takes the sample from the cervix using a speculum<br>2. Healthcare practitioner takes a vaginal swab sample<br>3. Self-taken vaginal swab sample<br>4. Urine sample | Categorical |
| Location to take the sample | Place where the sample will be taken | 1. Home<br>2. Healthcare setting | Categorical |
| Feeling of discomfort and/or pain | Feeling of discomfort or pain while taking the sample | 1. No discomfort or pain<br>2. Mild discomfort and/or pain<br>3. Moderate discomfort and/or pain<br>4. Severe discomfort and/or pain | Categorical |
| Contact with healthcare practitioner | Opportunity to talk with healthcare practitioner | 1. No contact<br>2. Contact via phone<br>3. Contact in person | Categorical |
| Frequency of screening | How often will the sample be taken for screening? | 1. Yearly<br>2. Every two years<br>3. Every three years<br>4. Every five years | Continuous |

In the time since the attribute selection process was conducted, a further study exploring women’s views of self-sampling for non-attenders has been published in the UK (42). This study involved women completing a postal questionnaire delivered alongside a self-testing kit as part of a clinical trial. As part of this questionnaire, free text comments were collated and analysed to identify key themes in responses. These included the ease of use of the test, the positive experience of self-sampling, issues with self-sampling, worries about test accuracy and self-confidence in test completion. The themes identified had been covered in the other studies identified in the review in this paper so no additional attributes were considered in the DCE design.

### Creating an Experimental Design

Experimental design is the process through which distinct combinations of attributes and their corresponding levels are transformed into choice sets in which participants express their preferences by making choices (43). An effective experimental design ensures that preferences for all of the attributes and levels can be estimated while minimising uncertainty in the results. When all possible combinations of attribute levels are included in the choice sets, this is known as full factorial design.

Taking the attributes and levels presented in Table 1, the full factorial for choice task A would give 1152 potential questions and 1,327,104 choice sets which is excessively large for practical use and might also allow occurrence of illogical combinations.

To reduce the number of choice sets to a manageable size to prevent placing excessive cognitive demands on the participants, D-efficient designs were generated using Ngene software (44) for choice task A and B. The d-efficient design of choice task A was designed with restrictions to prevent illogical combinations of attributes and levels from occurring together. For example, a healthcare practitioner would not be able to take the sample in a patient’s home, so these levels were prevented from occurring together.

A total of 24 questions were designed and divided into 3 blocks with 8 questions per block. The survey was designed to identify main effects only. At the end of the task A questions, 3 DCE tasks were included with fixed attributes representing different ways that cervical screening could be offered in practice. In the first of these, participants will be asked to choose between screening methods with levels representing current healthcare professional sampling or having no screening. In the second, they will be asked to choose between screening methods with levels representing vaginal self-sampling as it would likely be implemented in the NHS, compared to no screening. The final question will ask participants to choose between two sampling approaches representing current health professional or vaginal self-sampling compared to no screening. The aim of these questions is to determine whether uptake for screening is likely to be higher if a choice of sampling method is offered compared to a recommendation that everyone receives the same type of screen.

### Designing the Survey

The survey questionnaire will include five sections. The first section will include a participant information sheet explaining the study and will include a question that asks participants if they consent to taking part in the study. The next section will introduce the study, providing background information about the purpose of the study. The second section will offer detailed information on cervical cancer and different methods of collecting samples for cervical screening. This information will be designed using current NHS information leaflets for cervical screening and adapted to include information about self-sampling methods. The information will be checked for completeness and readability by members of a public advisory group (n=2). This information is intended to equip participants with the necessary knowledge to make informed decisions in the section following.

The third section will be the DCE questions where each participant will be asked to answer a series of choice questions. This will include 8 choice questions with varying attributes and levels and then 4 supplementary questions showing fixed scenarios: a realistic representation of healthcare-practitioner obtained cervical sampling versus no screening; a realistic representation of vaginal self-sampling versus no screening; and a choice of realistic representations of healthcare-practitioner obtained cervical sampling or vaginal self-sampling; and finally a choice of a realistic representation of healthcare-practitioner sampling, vaginal self-sampling, and urine self-sampling. Participants will be randomised to receive different blocks of questions. First, each attribute and corresponding levels will be described in detail and then the choice questions will be presented to the participants. The final section will gather additional sociodemographic information including: age; ethnicity; religion; employment status; education; disability; sexual orientation; gender; whether they have been sexually active in the past; whether they have been invited to cervical screening in the last 5 years; whether they have attended cervical screening in the last 5 years; awareness that screening utilises HPV testing; views about self-sampling; sources of health information; and degree of risk seeking behaviour. The full survey will be designed in the survey software Qualtrics and fielded online.

An example of choice question that will be presented in DCE section of the survey is presented in Table 3.

**Table 3.**
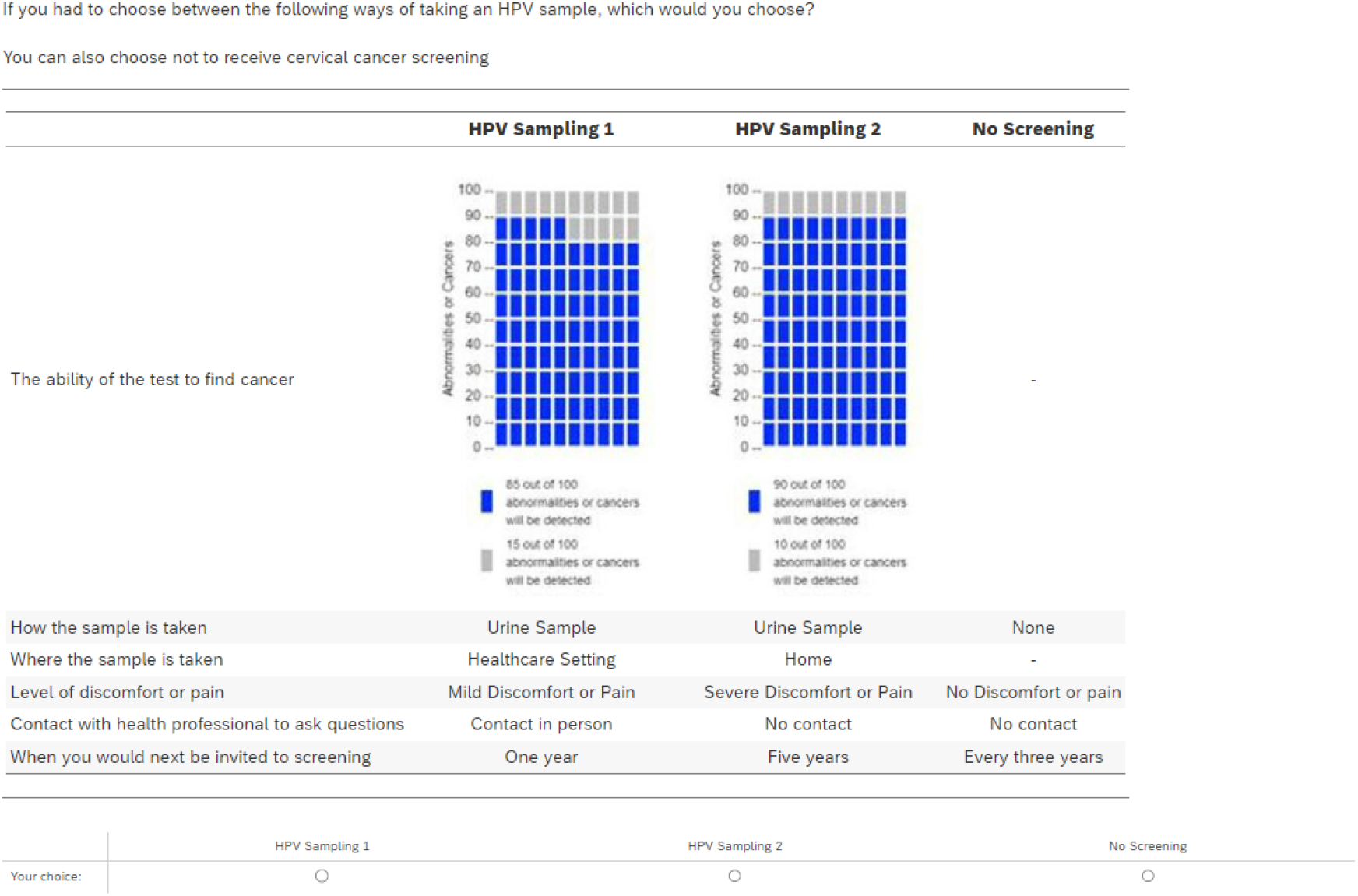
Example Choice Question^1^

|  | HPV Sampling 1 | HPV Sampling 2 | No Screening |
| --- | --- | --- | --- |
| The ability of the test to find cancer | 85 out of 100 abnormalities or cancers will be detected<br>15 out of 100 abnormalities or cancers will be detected | 90 out of 100 abnormalities or cancers will be detected<br>10 out of 100 abnormalities or cancers will be detected | - |
| How the sample is taken | Urine Sample | Urine Sample | None |
| Where the sample is taken | Healthcare Setting | Home | - |
| Level of discomfort or pain | Mild Discomfort or Pain | Severe Discomfort or Pain | No Discomfort or pain |
| Contact with health professional to ask questions | Contact in person | No contact | No contact |
| When you would next be invited to screening | One year | Five years | Every three years |

Your choice: ☐ HPV Sampling 1 ☐ HPV Sampling 2 ☐ No Screening

### Pilotingrnof the survey

Before fielding a discrete choice experiment in a full sample, piloting is required to test the survey. Piloting can improve the survey by checking participant understanding of the description of attributes and levels, length of survey, adequacy of design and data collection ensuring that the data can be optimally analysed.

To see if there is any adjustment needed to the final survey draft, a pre-pilot study will be conducted using face-to-face think aloud interviews with members of the public advisory group. During these interviews, participants will be asked to share their thought process while completing the survey. This approach allows researchers to understand the aspects of the survey where participants might misunderstand the questions or find them unclear (45). By applying the think aloud approach, researchers can identify potential issues and make necessary adjustments to improve the survey.

Following the pre-pilot, a quantitative pilot study will be conducted with a sample of 50 participants, including individuals from ethnically diverse backgrounds (such as Asian, Black, Caribbean, Arab). This pilot will check for design and coding errors, ensuring that all model coefficients can be estimated.

### Participant sampling and recruitment

There is no set rule on how many participants can be recruited for the DCE as estimating sample size for a DCE is quite challenging (17). The sample size depends on several factors such as the number of attributes and levels, choice tasks, continuous and categorical variables, and the degree of heterogeneity in responses. A sample of around 1000 is aimed for this study to increase the power of the study as well as ensure sufficient power for investigating heterogeneity of preferences in different groups.

The relevant study population will be individuals who have cervix and are between the ages of 25 to 64 years, and therefore eligible for cervical screening. The target population will be recruited by a commercial participant panel recruiter who will send a survey link to potential participants.

### Quality Control

It is necessary to ensure that the responses received from participants represent their true preferences as closely as possible. It is possible for DCE surveys to receive fraudulent responses, particularly when fielded online. Also, respondents may complete the survey quickly without fully reading all of the information or only focus on certain attributes and levels. Such responses when included in the analysis can result in biased results. Although econometric models account for random respondents, retaining these inaccurate responses can potentially create concern for the results of other survey questions.

To mitigate this risk, several quality control measures will be taken using tools in-built in Qualtrics: responders completing the survey too quickly (more than two standard deviations from the median) will be flagged, invisible Captcha technology will be used to help flag responses from bots, and a flag will be added to responses exhibiting potential straightlining (choosing the same option for all DCE tasks). Data analysis will be conducted with flagged responses included with additional sensitivity analysis conducted with flagged responses excluded.

### Analysis Approach

Data analysis will be conducted in R software using the Apollo package (46). Participants’ demographic information and responses to attitudinal questions will be summarised.⍰ The number of participants flagged as potentially fraudulent will be reported.⍰ The results of the DCE questions with fixed attributes for different realistic screening scenarios will be reported indicating the proportion of respondents who would choose healthcare professional sampling compared to no sampling, self-sampling compared to no sampling, healthcare professional or self-sampling compared to no sampling, and vaginal self-sampling compared to urine self-sampling and healthcare professional self-sampling.

For each DCE choice task, a process will be undertaken to select the functional form and model approach that best explains the data. A conditional logistic regression model will be estimated for each DCE with categorical variable effects coded, continuous variables assumed to be linear, and a single constant included to represent the probability of opting in versus opting out. Different model functional forms will be estimated whereby two constants are used to represent the probability of selecting hypothetical risk prediction or feedback scenario A or scenario B. This serves as a test as to whether participants were always choosing scenario A or B regardless of the levels shown. All tests of model specification will be made by comparing the Bayesian Information Criterion (BIC)⍰ of the different models. If a model specification is found to result in a lower BIC value then this suggests that the model specification adds sufficient additional explanatory power for the number of additional parameters in the model. ⍰

Analytical tests will be conducted to determine if there is evidence of non-linearity in the continuous variables (see Table 1) and y in the model. Each continuous variable will be coded in quadratic, log, and piece-wise forms to determine if any of these specifications reduces the BIC. Upon selection of the final functional form for the model, random parameter logit models will be estimated to determine if a model which allows for preference heterogeneity provides a better fit for the data. First an uncorrelated random parameter logit will be estimated for each DCE and a fully correlated random parameter logit will be estimated. The fully correlated model allows for differences in error between participants as well as differences in preferences.⍰

To investigate heterogeneity in preferences for sampling approach, latent class analysis will be used to identify groups of participants with similar preferences. The number of classes to include will be decided using an iterative approach comparing the BIC of each model as classes are added until the BIC no longer reduces. A further model will be fit to determine if any demographic variables are correlated with class membership.

Using the best model as determined using the BIC, the uptake for different potential sampling approaches for cervical screening will be estimated. The potential uptake for different approaches to sampling in the latent class groups will also be estimated.

### Dissemination

The results of the DCE will be published in a peer-reviewed journal. A Shiny package created using the software R will be used to make an app which will allow decision makers, such as the UK National Screening Committee, to combine different attributes and levels and explore the potential uptake for the resulting sampling approaches as predicted by the model. This will include uptake in the different latent class groups.

### Conclusion

This study has outlined the design of a discrete choice experiment (DCE) to explore the preferences for sampling method for cervical screening as part of the NHSCSP. The survey will now be piloted, fielded, and the data analysed to determine people’s preferences for different approaches to sampling methods for cervical screening and their potential uptake in clinical practice.

## Data Availability

There is no data associated with this article

## Footnotes

1 When the question is shown to participants the answer is automatically scaled so that the button for each answer is directly underneath the corresponding alternative

## REFERENCES

1. Cervical cancer [Internet]. [cited 2024 Mar 26]. Available from: https://www.who.int/news-room/fact-sheets/detail/cervical-cancer

2. Cancer Research UK [Internet]. 2015 [cited 2024 Mar 26]. Cervical cancer statistics. Available from: https://www.cancerresearchuk.org/health-professional/cancer-statistics/statistics-by-cancer-type/cervical-cancer

3. Bhatla N, Aoki D, Sharma DN, Sankaranarayanan R. Cancer of the cervix uteri: 2021 update. Int J Gynecol Obstet. 2021;155(S1):28–44.

4. Causes | Background information | Cervical cancer and HPV | CKS | NICE [Internet]. [cited 2024 Mar 26]. Available from: <https://cks.nice.org.uk/topics/cervical-cancer-hpv/background-information/causes/>

5. Marth C, Landoni F, Mahner S, McCormack M, Gonzalez-Martin A, Colombo N, et al. Cervical cancer: ESMO Clinical Practice Guidelines for diagnosis, treatment and follow-up. Ann Oncol Off J Eur Soc Med Oncol. 2017 Jul 1;28(Suppl_4):iv72–83.

6. NHS England. NHS urges more women to take up cervical screening invitations [Internet]. 2024 [cited 2024 Oct 7]. Available from: https://www.england.nhs.uk/2024/06/nhs-urges-more-women-to-take-up-cervical-screening-invitations/

7. NHS England Digital [Internet]. [cited 2024 Mar 26]. Cervical Screening Programme, England - 2022-2023 [NS]. Available from: <https://digital.nhs.uk/data-and-information/publications/statistical/cervical-screening-annual/england-2022-2023>

8. Landy R, Pesola F, Castañón A, Sasieni P. Impact of cervical screening on cervical cancer mortality: estimation using stage-specific results from a nested case–control study. Br J Cancer. 2016 Oct;115(9):1140–6.

9. Bennett KF, Waller J, Chorley AJ, Ferrer RA, Haddrell JB, Marlow LA. Barriers to cervical screening and interest in self-sampling among women who actively decline screening. J Med Screen. 2018 Dec;25(4):211–7.

10. Marlow LAV, Waller J, Wardle J. Barriers to cervical cancer screening among ethnic minority women: a qualitative study. J Fam Plann Reprod Health Care. 2015 Oct 1;41(4):248–54.

11. Davies-Oliveira JC, Round T, Crosbie EJ. Cervical screening: the evolving landscape. Br J Gen Pract. 2022 Aug 1;72(721):364–5.

12. Moser K, Patnick J, Beral V. Inequalities in reported use of breast and cervical screening in Great Britain: analysis of cross sectional survey data. BMJ. 2009 Jun 16;338:b2025.

13. RIVM. Information about cervical cancer screening. 2024 [cited 2025 Jan 30]. Information about cervical cancer screening. Available from: https://www.rivm.nl/en/cervical-cancer-screening-programme/information-materials

14. Woo YL, Khoo SP, Gravitt P, Hawkes D, Rajasuriar R, Saville M. The Implementation of a Primary HPV Self-Testing Cervical Screening Program in Malaysia through Program ROSE-Lessons Learnt and Moving Forward. Curr Oncol Tor Ont. 2022 Oct 2;29(10):7379–87.

15. Ylli A, Filipi K, Shundi L, Fico A, Risto M, Xhani A. New cervical screening program in Albania. Access and barriers in all levels of health system. Eur J Public Health. 2020 Sep 1;30(Supplement_5):ckaa166.487.

16. Serrano B, Ibáñez R, Robles C, Peremiquel-Trillas P, de Sanjosé S, Bruni L. Worldwide use of HPV self-sampling for cervical cancer screening. Prev Med. 2022 Jan 1;154:106900.

17. HPValidate cervical screening self-sampling study nears completion – UK National Screening Committee [Internet]. 2023 [cited 2024 Mar 27]. Available from: https://nationalscreening.blog.gov.uk/2023/06/21/hpvalidate-cervical-screening-self-sampling-study-nears-completion/

18. NHS England⍰ » NHS gives women Human Papillomavirus Virus (HPV) home testing kits to cut cancer deaths [Internet]. [cited 2024 Mar 27]. Available from: https://www.england.nhs.uk/2021/02/nhs-gives-women-hpv-home-testing-kits-to-cut-cancer-deaths/

19. Cancer Research UK [Internet]. 2022 [cited 2024 Mar 27]. A study looking at using urine to screen for cervical cancer (ACES). Available from: https://www.cancerresearchuk.org/about-cancer/find-a-clinical-trial/a-study-looking-at-using-urine-to-screen-for-cervical-cancer-aces

20. Arbyn M, Verdoodt F, Snijders PJF, Verhoef VMJ, Suonio E, Dillner L, et al. Accuracy of human papillomavirus testing on self-collected versus clinician-collected samples: a meta-analysis. Lancet Oncol. 2014 Feb 1;15(2):172–83.

21. Mathews C, Brentnall A, Rebolj M, Sargent A, Cuschieri K, Denton K. HPValidate: clinical validation of hrHPV test system using self-collected vaginal samples in NHS England commissioned laboratories providing cervical screening services [Internet]. Queen Mary University of London; 2024 Sep [cited 2025 Feb 12]. Available from: https://www.qmul.ac.uk/fmd/media/smd/documents/research/hpv-self-collection-test-accuracy-report-hpvalidate-lot1.pdf

22. Tatara T, Wnuk K, Miazga W, Świtalski J, Karauda D, Mularczyk-Tomczewska P, et al. The Influence of Vaginal HPV Self-Sampling on the Efficacy of Populational Screening for Cervical Cancer—An Umbrella Review. Cancers. 2022 Nov 30;14(23):5913.

23. Aitken CA, Inturrisi F, Kaljouw S, Nieboer D, Siebers AG, Melchers WJG, et al. Sociodemographic Characteristics and Screening Outcomes of Women Preferring Self-Sampling in the Dutch Cervical Cancer Screening Programme: A Population-Based Study. Cancer Epidemiol Biomark Prev Publ Am Assoc Cancer Res Cosponsored Am Soc Prev Oncol. 2023 Feb 6;32(2):183–92.

24. Davies JC, Sargent A, Pinggera E, Carter S, Gilham C, Sasieni P, et al. Urine high-risk human papillomavirus testing as an alternative to routine cervical screening: A comparative diagnostic accuracy study of two urine collection devices using a randomised study design trial. BJOG Int J Obstet Gynaecol. 2024 Oct;131(11):1456–64.

25. Van Keer S, Peeters E, Vanden Broeck D, De Sutter P, Donders G, Doyen J, et al. Clinical and analytical evaluation of the RealTime High Risk HPV assay in Colli-Pee collected first-void urine using the VALHUDES protocol. Gynecol Oncol. 2021 Sep;162(3):575–83.

26. Nishimura H, Yeh PT, Oguntade H, Kennedy CE, Narasimhan M. HPV self-sampling for cervical cancer screening: a systematic review of values and preferences. BMJ Glob Health. 2021 May;6(5):e003743.

27. Rebolj M, Sargent A, Njor SH, Cuschieri K. Widening the offer of human papillomavirus self-sampling to all women eligible for cervical screening: Make haste slowly. Int J Cancer. 2023 Jul 1;153(1):8–19.

28. Vass CM, Rigby D, Payne K. Investigating the Heterogeneity in Women’s Preferences for Breast Screening: Does the Communication of Risk Matter? Value Health. 2018 Feb;21(2):219–28.

29. de Bekker-Grob EW, Ryan M, Gerard K. Discrete choice experiments in health economics: a review of the literature. Health Econ. 2012;21(2):145–72.

30. Lancsar E, Louviere J. Conducting discrete choice experiments to inform healthcare decision making: a user’s guide. PharmacoEconomics. 2008;26(8):661–77.

31. Bridges JFP, Hauber AB, Marshall D, Lloyd A, Prosser LA, Regier DA, et al. Conjoint analysis applications in health--a checklist: a report of the ISPOR Good Research Practices for Conjoint Analysis Task Force. Value Health J Int Soc Pharmacoeconomics Outcomes Res. 2011 Jun;14(4):403– 13.

32. A Reporting Checklist for Discrete Choice Experiments in Health: The DIRECT Checklist | PharmacoEconomics [Internet]. [cited 2024 Sep 10]. Available from: https://link.springer.com/article/10.1007/s40273-024-01431-6

33. A New Approach to Consumer Theory | Journal of Political Economy: Vol 74, No 2 [Internet]. [cited 2024 Sep 10]. Available from: https://www.journals.uchicago.edu/doi/10.1086/259131

34. Coast J, Horrocks S. Developing attributes and levels for discrete choice experiments using qualitative methods. J Health Serv Res Policy. 2007 Jan;12(1):25–30.

35. Waller J, Mccaffery K, Forrest S, Szarewski A, Cadman L, Austin J, et al. Acceptability of unsupervised HPV self-sampling using written instructions. J Med Screen. 2006 Dec;13(4):208–13.

36. Cadman L, Ashdown-Barr L, Waller J, Szarewski A. Attitudes towards cytology and human papillomavirus self-sample collection for cervical screening among Hindu women in London, UK: a mixed methods study. J Fam Plann Reprod Health Care. 2015 Jan;41(1):38–47.

37. Szarewski A, Cadman L, Ashdown-Barr L, Waller J. Exploring the acceptability of two self-sampling devices for human papillomavirus testing in the cervical screening context: a qualitative study of Muslim women in London. J Med Screen. 2009;16(4):193–8.

38. Robb KA. The integrated screening action model (I-SAM): A theory-based approach to inform intervention development. Prev Med Rep. 2021 Sep 1;23:101427.

39. Bansback N, Hole AR, Mulhern B, Tsuchiya A. Testing a discrete choice experiment including duration to value health states for large descriptive systems: Addressing design and sampling issues. Soc Sci Med. 2014;114:38–48.

40. Bech M, Kjaer T, Lauridsen J. Does the number of choice sets matter? Results from a web survey applying a discrete choice experiment. Health Econ. 2011 Mar;20(3):273–86.

41. Soekhai V, de Bekker-Grob EW, Ellis AR, Vass CM. Discrete Choice Experiments in Health Economics: Past, Present and Future. PharmacoEconomics. 2018;1–26.

42. Drysdale H, Marlow LAV, Lim A, Waller J. Experiences of Self-Sampling and Future Screening Preferences in Non-Attenders Who Returned an HPV Vaginal Self-Sample in the YouScreen Study: Findings From a Cross-Sectional Questionnaire. Health Expect Int J Public Particip Health Care Health Policy. 2024 Aug;27(4):e14118.

43. Reed Johnson F, Lancsar E, Marshall D, Kilambi V, Mühlbacher A, Regier DA, et al. Constructing Experimental Designs for Discrete-Choice Experiments: Report of the ISPOR Conjoint Analysis Experimental Design Good Research Practices Task Force. Value Health. 2013 Jan 1;16(1):3– 13.

44. Ngene 1.4 [Internet]. Sydney, Australia: Choice Metrics; Available from: https://www.choice-metrics.com/

45. Ryan M, Watson V, Entwistle V. Rationalising the ‘irrational’: a think aloud study of discrete choice experiment responses. Health Econ. 2009 Mar;18(3):321–36.

46. Hess S, Palma D. Apollo: A flexible, powerful and customisable freeware package for choice model estimation and application. J Choice Model [Internet]. 2019 [cited 2024 Sep 10];32(C). Available from: https://econpapers.repec.org/article/eeeeejocm/v_3a32_3ay_3a2019_3ai_3ac_3a4.htm

