## Supplementary materials for "Understanding preferences for self-sampling in a national cervical screening programme: a protocol for a discrete choice experiment"

Supplementary appendix 1

Search Strategy

Ovid MEDLINE(R) <1946 to January Week 5 2024>

1	qualitative studies.mp. or Qualitative Research/

2	Focus Groups/

3	Interview/

4	Observation/

5	ethnography.mp.

6	Grounded Theory/

7	acceptability.mp.

8	Health Knowledge, Attitudes, Practice/ or facilitators.mp.

9	barriers.mp.

10	challenge*.mp.

11	preference*.mp.

12	acceptance.mp.

13	facilitate*.mp.

14	self sampl*.mp.

15	self collect*.mp.

16	self administered.mp.

17	self test*.mp.

18	self evaluation.mp. or Diagnostic Self Evaluation/

19	HPV.mp. or Papillomavirus Infections/ or Uterine Cervical Neoplasms/

20	cervical.mp.

21	cervix.mp. or Cervix Uteri/

22	cervical cancer.mp. or Uterine Cervical Neoplasms/

23	1 or 2 or 3 or 4 or 5 or 6	153348

24	7 or 8 or 9 or 10 or 11 or 12 or 13

25	14 or 15 or 16 or 17 or 18

26	19 or 20 or 21 or 22

27	23 and 24 and 25 and 26

Embase <1974 to 2024 Week 06>

1	qualitative studies.mp. or qualitative research/

2	structured interview/ or telephone interview/ or audio interview/ or interview.mp. or unstructured interview/ or video interview/ or semi structured interview/ or interview/

3	participant observation/ or observation/

4	ethnography/ or ethnology/

5	grounded theory/

6	acceptability.mp. or program acceptability/

7	facilitators.mp.

8	barriers.mp.

9	challenge*.mp.

10	preference*.mp. or social preference/ or patient preference/

11	acceptance.mp.

12	facilitate*.mp.

13	sampling/ or self sampl*.mp.

14	self collect*.mp.

15	self administered.mp.

16	self-testing/ or self test.mp.

17	self evaluation/

18	HPV.mp. or Wart virus/

19	Papillomavirus.mp. or Papillomaviridae/

20	uterine cervix cancer/ or cancer screening/

21	uterine cervix/

22	1 or 2 or 3 or 4 or 5

23	6 or 7 or 8 or 9 or 10 or 11 or 12

24	13 or 14 or 15 or 16 or 17

25	18 or 19 or 20 or 21

26	22 and 23 and 24 and 25

**Supplementary materials 2: Attribute identification**

**Supplementary Table 2.1 Long list of attributes derived from literature revie**

| **Cultural aspect** | **Device itself** | **Social** | **Communication** | **Knowledge aspect** | **Trans man/non-binary** | **Others** |
| --- | --- | --- | --- | --- | --- | --- |
| Sexually inactivity (including widows, unmarried women etc) believing that they do not need screening | Reliability of the test | Younger generation versus older generation – older generation reluctant to use | Testimonials from other women in the information sheet | Link between HPV positivity and cervical cancer | Genital dysphoria – would increase by having to engage with own genitalia | Ordering kit requires action - barrier to people with language, disability |
| Fear of losing virginity | Size of the sampling kit – small preferred | Discuss with other women and partners | Tailored information with explicit consequences of late detection in invitation letter | The test is for HPV and cervical sample will be needed if the result is HPV positive | Genital dysphoria – would decrease by not having to expose their genitals to someone else | Anxiety of the result |
| Associated with sexual activity before marriage | Confidence in doing the screening/Fear of not doing it correctly |  | Invitation letter – upfront information on test accuracy |  | Accessibility | Flexible option of returning the kit |
| Unmarried women living with family might have dilemma to have the kit sent to their home address | Instructions that come with sampling method |  | More information in instruction leaflet – how to store the sample and post (eg. posting the sample on a hot day), timeframe to return the sample, reminder to post |  |  | May not fit in the mailbox, have to go to the post office |
| Privacy of home | Unfamiliar with the kit |  | Format of receiving result |  |  | Fear of detecting cancer |
| **Cultural aspect** | **Device itself** | **Social** | **Communication** | **Knowledge aspect** | **Trans man/non-binary** | **Others** |
| Alleviates embarrassment | Not having someone to reassure them that they are using the kit properly |  | Fear of post office losing the sample and having to go through doing the test again |  |  |  |
| Only having one sexual partner mean you can’t get HPV/cervical cancer | Fear of increasing infection – if the device is lost inside the body |  |  |  |  |  |
| Body shyness, unwillingness to touch or engage with own genitals | Risk of hurting oneself if not done correctly |  |  |  |  |  |
|  | Fear of not collecting enough sample |  |  |  |  |  |
|  | Fear of doing the sampling wrong and getting the wrong result |  |  |  |  |  |
|  | Incorrect belief that they may not able to collect sample properly as they might not reach their cervix with the swab |  |  |  |  |  |
|  | Sterilisation of the swab |  |  |  |  |  |
|  | First time maybe shown by health professional |  |  |  |  |  |
|  | Fear of missing cancer |  |  |  |  |  |
